# Experienced hearing aid users’ perspectives of assessment and communication within audiology: A qualitative study using digital methods

**DOI:** 10.1101/2021.09.08.21263074

**Authors:** Bhavisha Parmar, Kinjal Mehta, Deborah Vickers, Jennifer K. Bizley

## Abstract

**Objective:** To explore experienced hearing aid users’ perspectives of audiological assessments and the patient-audiologist communication dynamic during clinical interactions.

**Design:** A qualitative study was implemented incorporating both an online focus group and online semi-structured interviews. Sessions were audio-recorded and transcribed verbatim. Iterative-inductive thematic analysis was carried out to identify themes related to assessment and communication within audiology practice.

**Study samples:** Seven experienced hearing aid users took part in an online focus group and 14 adults participated in semi-structured interviews (age range: 22 - 86 years; 9 males, 11 females).

**Results:** Themes related to assessment included the unaided and aided testing procedure and relating tests to real world hearing difficulties. Themes related to communication included the importance of communication strategies, explanation of test results and patient centred care in audiology.

**Conclusion:** To ensure that hearing aid services meet the needs of the service users, we should explore user perspectives and proactively adapt service delivery. This approach should be ongoing, in response to advances in hearing aid technology. Within audiology, experienced hearing aid users’ value 1) comprehensive, relatable hearing assessment, 2) clear, concise, deaf aware patient-audiologist communication, 3) accessible services and 4) a personalised approach to recommend suitable technology and address patient specific aspects of hearing loss.

## Introduction

Hearing loss is a growing public health concern that negatively impacts daily life including communication, behavioural and social interaction and emotions, identity, and psychological wellbeing (Vas et al., 2017; Wilson et al., 2017). The benefits of hearing aids, for people with hearing loss, are well established (Acar et al., 2011; Chisolm et al., 2007; Ferguson et al., 2017; Hornsby, 2013; Humes et al., 2002). However, hearing aid acceptance and uptake remains low worldwide and self-reported benefits vary (Bisgaard & Ruf, 2017; Chien & Lin, 2012; Humes et al., 2002; Lopez-Poveda et al., 2017).

Effective patient-clinician communication is an essential part of patient-centred care (Zill et al., 2015); however, clinicians working with people with hearing loss may face additional challenges due to the impact of hearing loss on communication (Mick et al., 2014). Within audiology, the patient-audiologist interaction has been recognised as an important factor contributing to patient satisfaction, hearing aid adoption and hearing aid use (Grenness et al., 2014; Ismail et al., 2019; Poost-Foroosh et al., 2011; Sciacca et al., 2017), and has been investigated across a variety of populations and settings (Ekberg et al., 2014b; Mehta et al., 2019; Tai et al., 2019; Watermeyer et al., 2020; Watermeyer et al., 2017). Importantly, research demonstrates that audiologists tend to use clinician-centred approaches in practice, without addressing patients’ concerns (Tai et al., 2019), in stark contrast to the patient-centred approach at the core of audiologists’ clinical responsibility (American Speech-Language-Hearing Association, 2018). Furthermore, there is evidence to suggest that psychosocial support is not routinely provided in audiology practice (Bennett et al., 2020; Ekberg et al., 2014a). Audiologists vary considerably in their interaction style with patients, in some cases they use overly-detailed scientific explanations without personalised care (Watermeyer et al., 2017).

A patient’s hearing aid usage and communication needs may differ based on factors including the degree of hearing loss (Hartley et al., 2010), lifestyle (Barker et al., 2016), perceived benefit of hearing aids, perceived hearing disability, shared decision making and the role of communication partners (Hickson et al., 2014; Meyer et al., 2014; Ng & Loke, 2015). It is also important to recognize that the provision of audiology-based healthcare services varies considerably around the world, and, in some cases, hearing aid users may have access to different service providers. An international investigation of hearing help seeking and rehabilitation revealed mixed reviews regarding the hearing assessment process and the applicability of test results to everyday listening difficulties (Laplante-Lévesque et al., 2012), highlighting the need for specific patient-centred approaches in this area to help professionals improve communication. Furthermore, some hearing aid users had preconceived notions about private versus public hearing healthcare. They reported that relational competence, including the clinicians’ overall communication manner, was an important component of trust within the patient-audiologist dynamic (Preminger et al., 2015).

Recently, research has focussed on enhancing the ecological validity of audiological assessment (Keidser et al., 2020) and the development of tools to effectively explain assessment results to patients (Klyn et al., 2019), with the view of individualising care. Considering these findings, the influence of the patient-audiologist interaction, and the importance of ensuring hearing aid services meet the needs of the service users, we should explore user perspectives and proactively adapt service delivery. This study investigates experienced hearing aid users’ perspectives of audiological assessments using iterative-inductive thematic analysis of semi-structured interviews and an online focus group. Interviews focused on the style of communication during patient-audiologist interactions, including the type of information given to patients by audiologists.

## Methods

### Ethical Approval

This study was approved by the UCL Ethics Committee (project no. 3866/001). Data was kept in compliance with the General Data Protection Regulation (EU 2016/679). Each participant provided written consent, personal identities were anonymised during transcription and audio recordings were subsequently destroyed.

### Recruitment

Experienced hearing aid users (i.e., adults with over four years’ experience of regular hearing aid use) with no known neurological conditions were identified through a database of individuals with hearing loss who had volunteered to take part in research studies at the UCL Ear Institute, London, England. Experienced hearing aid users were targeted for this study (rather than first time users) because of their increased exposure to a range of audiology services and clinicians as well as increased awareness of the hearing healthcare pathway. Email invitations were sent to potential participants who had been in contact with their audiology provider within the last 24 months.

Purposive sampling with maximum variation was employed to prioritize enrolment of eligible participants in the one-to-one interviews to maximise diversity of views and experiences (Suri, 2011). Upon expressed interested, participants completed a demographic questionnaire adapted from Dawes et al (2014). A sampling matrix was then employed to ensure interviewees with diverse backgrounds were included (see Supplementary Material).

### Participants

A total of 21 adults with an average of 17 years of hearing aid use participated in this study with different populations for the focus group (7 participants) and for the semi-structured interviews (14 participants). Demographics are provided in Table 1 (see Supplementary Material for individual demographics).

**Table 1:** Summary of participant demographics

|  | Characteristics | Frequency |
| --- | --- | --- |
| Age group | 18-30 | 1 |
|  | 31-40 | 1 |
|  | 41-50 | 2 |
|  | 51-60 | 6 |
|  | 61-70 | 6 |
|  | 71-80 | 2 |
|  | 81+ | 3 |
| Sex | *Female* | 12 |
|  | *Male* | 9 |
| Level of education | *Secondary school* | 4 |
|  | *Trade qualification* | 2 |
|  | *Diploma* | 2 |
|  | *University degree* | 7 |
|  | *Post graduate qualification* | 5 |
|  | *PhD* | 1 |
| Work status | *Working* | 11 |
|  | *Retired* | 10 |
| Audiology provider | *Private* | 9 |
|  | *Public* | 12 |

### Procedure

#### Topic Guide

The present study implemented a semi-structured approach in an effort to understand “participants’ experiences, how they describe their experiences, and the meaning they make of those experiences” in an extended conversational format (Rubin, 2012). The topic guide was developed based on open-ended questions (e.g., “Describe your experiences of hearing assessment”) and follow-up probes (see Supplementary Material for the complete topic guide). Once compiled, third-party audiologists and experienced hearing aid users reviewed the topic guide content and interpretability (see Figure 1).

**Figure 1.**
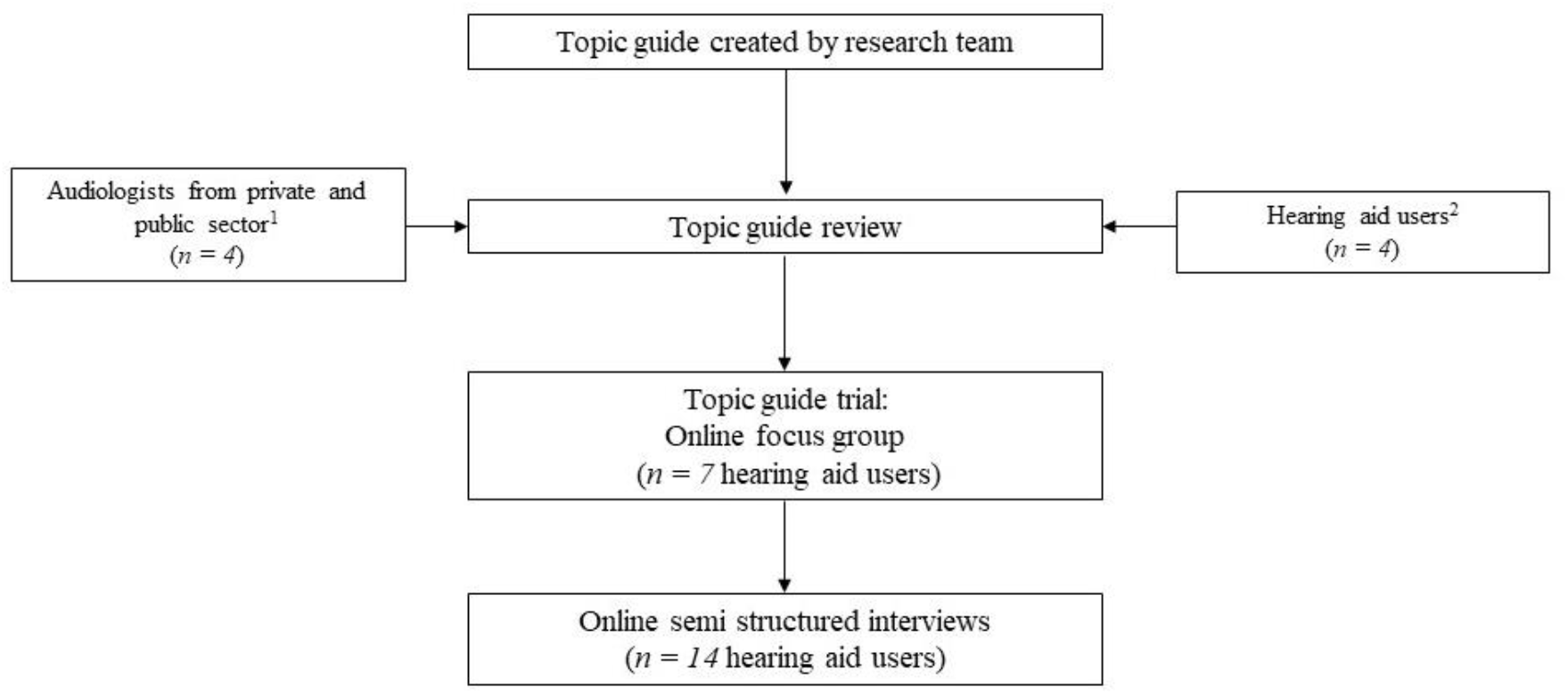
Flow chart detailing the progression from topic guide development to data collection. ^1^Audiologists suggested the inclusion of questions regarding the use of aided assessment methods, ^2^Hearing aid users recommended questions about accessibility and communication experiences within the audiology reception area/waiting area.

The focus group was carried out first, as an explorative process, to ensure the target discussion topics were easily interpretable, aligned with the main research questions, and were appropriate for use within the semi-structured interview format (Cyr, 2016). All focus group participants reported good interpretability of questions and no changes were required prior to inclusion in the semi-structured interviews. Thus, the focus group data was included in the present analysis and the topic guide used in the focus group was used for all interviews.

#### Online Protocol

Focus group and interview sessions were conducted between January and March 2021. The focus group was carried out via Zoom because all participants had previous experience with this platform, did not require closed captions, and were comfortable with the visual gallery layout. Semi-structured interview participants chose their preferred online platform (Zoom or Microsoft Teams) based on their personal communication needs (e.g., closed captioning). All participants were offered a 1:1 trial call prior to the main session to check audio and internet quality.

The interview facilitator (B.P.) hosted each video call from a professional setting within their own home. Participants took part in the online video conference from their own homes and were encouraged to sit in a well-lit room with minimal background noise. Participants wore their hearing aids throughout the video call and chose to use either headsets, Bluetooth streamers, or loudspeakers based on their communication needs. The virtual waiting room was open an hour before each call was scheduled to allow for audio and/or Bluetooth connection testing.

#### Focus group

Focus groups of six to ten participants are generally optimal for ensuring all individuals are able to contribute and provide a range of views without being led by the consensus of a larger group (Morgan, 1998). The nature of the online, video conference environment also impacts group facilitation and communication considerably, particularly for people with hearing difficulties (Dawes et al., 2014). Thus, the focus group was limited to seven participants and was 90 minutes in duration. Participants were encouraged to use a laptop or tablet (rather than a smartphone) and to keep their cameras switched on in gallery view to optimise audibility and access the non-verbal cues.

Specific instructions were provided to respondents prior to the focus group session time to optimise communication and replicate the interactive nature of in-person focus groups as best as possible (see Supplementary Material). During the session, all participants’ microphones were muted. Participants were able to unmute if they wanted to join the discussion; focus group members were asked to raise their hand (either physically or by using a virtual signalling feature within the video conferencing software), to indicate they would like to contribute to the discussion.

#### Semi-structured interviews

Due to the one-to-one nature of the semi-structured interview, participants were able to use their preferred communication device (including smartphones); all parties kept their cameras and microphones switched on for the duration of the interview. On average, each interview session took approximately 46 minutes (range: 35 - 58 minutes). To avoid fatigue, the facilitator ensured interviews did not exceed the maximum allotted time of one hour (Adams, 2015).

#### Data Analysis

Iterative-inductive thematic analysis with a descriptive phenomenological approach was used to identify, analyse and report patterns within the data (Braun & Clarke, 2006). The present thematic analysis uses a descriptive approach with a focus on lived experience.

Inductive (bottom up) thematic analysis was implemented by the first two authors, such that coded data were organised into general themes centring around experiences during audiology appointments and patient-audiologist communication. A saturation point was reached after inductive coding of the 11th interview (participant PQ) and was confirmed by carrying out three additional interviews for which no new themes emerged. The first and second authors double coded 25% of transcripts and any discrepancies were resolved at two separate timepoints within the study.

## Results

### Experience of hearing assessment

Here, participants focussed on their recent audiology hearing assessment experiences. Two key themes were identified: 1) testing procedure and 2) relating tests to real world hearing difficulties.

### Testing procedure

Participants described their experiences undergoing pure tone audiometry, including aspects of the patient-audiologist interaction and certain testing instructions that helped them feel more comfortable.

> *“Something that really stood out in my mind, no matter how faint the noise is that it’s just as important as the louder ones. Now in the past I remember I’ve sat there thinking can I hear that? … this time I was really determined that I was gonna press it if it was there was even a quiet sound”* – LK (age range: 51-55 years, male, private sector)

Some participants mentioned completing speech testing during audiology assessments and several reported the value for understanding the benefits of their hearing aids when switching between aided and unaided testing.

> *“It made me realise what I couldn’t hear, because he did a test first of all without my hearing aids in and then he read the words again in a different order with my hearing aids in and it made me understand what letter sounds that I couldn’t hear, what I was mixing up. That was very useful from a self-awareness point of view”* -HG (age range: 51-55 years, female, private & public sector)
>
> *And then I went private and he kind of explained to me, this is how it works in terms of picking up speech and really emphasised on speech in testing. And I was able to really kind of understand what I couldn’t hear and why I couldn’t hear certain sounds in relation to my hearing loss. So, it’s really interesting*. – RS (age range 35-39 years, female, private sector)

### Relating tests to real world hearing difficulties

Participants described the experience of aided assessment once new hearing aids had been fitted or existing hearing aids had been adjusted, with a variety of aided testing techniques e.g., speech perception testing or presentation of distractor stimuli. However, some participants reported that the audiologist tended to ask them for their opinions of the hearing aid sound quality rather than performing any aided assessments or using outcome measures.

> *“They ask: ‘how does that sound?’ & ‘are you comfortable with that?’ and yeah that’s usually how it’s done” -* WX (age range: 56-60, male, public sector)

The benefits and challenges of replicating externally valid situations in a controlled clinical environment were discussed in detail. For example, the significant limitation of conducting hearing aid fittings in a quiet room was emphasized, as was the benefit of using speakers to play background noise to simulate noisy environments. When realistic sounds and background noise were used within the audiology assessment, participants reported feeling more confident wearing the hearing aids outside of the clinic and expressed a lower need to return to the clinic for adjustments.

> *“And I think it gives you confidence to know that you can pick up certain speech with the hearing aids you’ve got, kind of the reassurance to know I can still pick up speech”* -RS (age range: 35-39 years, female, private sector)

Other respondents did indicate, however, that a simulated environment was still very unrealistic compared to everyday challenging listening situations.

> *“They would test different noises and then I’ll put one* [hearing aid] *in and the other one in to see how it felt and their voice as they were speaking to me, but it’s not a good test of the real world to be honest because of course you know my perfect world isn’t one of being quiet without background noise. That’s not how we hear”* - ML (age range: 40-45 years, female, public sector)

Some participants believed that it would be beneficial to record their listening difficulties in environments they experience regularly (e.g., home, work) or have a trial period wearing hearing aids outside the clinic with the option of providing immediate feedback.

> *“I can hear you in a completely silent room, but as soon I walk out the door it isn’t right. So, the advantage and it’s not perfect, but you can actually have the audiologist or receptionist or someone random person that happens to be in the building, come in and they can have a conversation while you’ve got music or street sounds on, so it gives you a much better idea because they can tweak it while you’re there”* -ON (age range: 56-60 years, female, private sector)
>
> *“I’ve had the thing that mimics restaurants. If only they could come with me or if only they could just come with me and tell me about the street, or come to work with me*.” - LK (age range: 51-55 years, male, private sector)
>
> *“You’ve got all the other kind of speakers that generate all sorts of background noise so that you can try them in background noise and then you can wander out in the street in them for a few minutes and come back” -* NO (age range: 56-60, private & public sector)

### Patient-Audiologist Communication

Participants discussed their experiences within specific scenarios they had experienced or in relation to communication techniques they hoped audiologists would incorporate in future meetings. Three key themes were identified: 1) importance of communication strategies, 2) explanation of test results, and 3) patient-centred care. Participants felt that the lack of individualised care and inadequate communication strategies amongst practitioners negatively impacted their experiences.

### Importance of communication strategies

Firstly, participants emphasized the importance of professional development training regarding communication skills and deaf awareness for all audiology staff, including receptionists. Participants also recounted specific situations where audiologists had not used adequate communication skills (e.g., conducting clinical questionnaires when the patient was not wearing their hearing aids) and indicated that some healthcare professionals did not seem aware of the communication barriers people with hearing aids face.

> *“I’ve had it where I’ve had my impressions taken, they’ve filled my ears obviously with the putty and start talking to me and I’ve not looked at them. And then it’s kind of… I laugh about it. Quite often I’m one of those I kind of brush things off quite a lot” -*QP (age range: 20-24 years, female, public sector)
>
> *“I mean many times I’ve been sat there unaided, with the audiologist talking to me, turning around to put the tube in and I think don’t you know… you of all people must know I can’t hear a thing you’re saying. It’s just annoying*.*” -*WX (age range: 56-60 years, male, public sector)
>
> *“Some audiologists really have… very minimal experience of working with patients like myself, with the type of hearing loss that I have. One had a tendency to over-pronounce and I said you don’t need to speak to me like I’m a five-year-old” -*RS (age range: 35-39 years, female, private sector)
>
> *“And the audiologist is clearly checking setting your hearing aids against whatever’s on the computer. And if you felt like they were concentrating on something, and then you had a question… Would you feel comfortable to ask it?” -*AB (age range: 81-85 years, male, public sector)
>
> To better understand the first-hand experience of hearing loss and the perspective of their patients, experienced hearing aid users suggested audiologists should spend time with temporary simulated hearing loss (e.g., ear plugs). The importance of having working loop systems and visual displays in the reception areas to alert patients was also stressed
>
> *“the first thing I would do is I would arrange for them to have a week with earplugs in. Just so they would appreciate what people who can’t hear are up against. It happens every time when I don’t have the aids in my ears because they’re adjusting them and they try and speak to me, I do that* [points to ear], *and they realise I can’t hear them*”-WX (age range: 56-60 years, male, public sector)
>
> *“Somebody with really good deaf awareness that faces you when they speak, they explain what they’re going to do before they do it, they’re not somebody that just comes and says right sit in that booth, I’ll put these on, press the button when you hear something”* – HG (age range: 51-55 years, female, private & public sector)

Finally, some participants reported positive experiences involving being seen by audiologists with hearing loss. In these cases, they felt the audiologist had an enhanced awareness of the impact of hearing loss and a unique ability to relate to the hearing difficulties experienced by patients.

> *“I think possibly if more deaf people were involved in audiology, not just one deaf audiologist, but if more people, more deaf people were involved all the way through then there might be some changes*.*” –* FE (age range: 61-65 years, female, public sector)
>
> *“My audiologist was Deaf and it was quite an experience. I knew that the person had really good understanding, really good deaf awareness because not all audiologists that I’ve seen have a good deaf awareness. -* HG (age range: 51-55 years, female, private & public sector)
>
> Some participants also reported communication barriers in the waiting area and with administrative staff.
>
> *“In the reception I feel like…do I really have to tell you that I’m deaf? I would’ve thought in this environment this is somewhere where I’m safe, you know where I’m with like-minded people and I don’t have to explain myself”* -LK (age range: 51-55 years, male, private sector)
>
> *“There’s a big waiting room, the audiologist calls my number and then they walked away and didn’t have a chance to see the lip reading or anything*.*”* -LM (age range: 71-74 years, male, public sector)

### Explanation of test results

Participants explained their general understanding of audiometry results and they reported engaging in varying levels of explanation with their audiologist when interpreting test results. Some respondents felt their audiologist’s description was sufficient for their needs and trusted the audiologist’s judgement without requiring additional detail. Others, however, were not shown the audiogram and felt they would have benefitted from more information., Some participants felt audiologists assumed audiograms had been explained to the patient in depth at an earlier date because they were long-term hearing aid users and thus were simply comfortable reporting when there were no significant changes to hearing thresholds, even if the patient was unfamiliar with audiogram reports.

> *“I mean it appears on the screen and I sort of look vaguely at the screen but not with great interest, I trust, I trust her*.*”* – TU (age range: 81-85 years, female, private sector)
>
> *“My audiologist just said something like, well there’s not much change*.*”* – CD (age range: 56-60 years, male, public sector)
>
> *“Because when they do the testing, they didn’t tell me the results of the testing, of how beneficial my hearing aids actually were. I just felt like I had this massive gap in my knowledge and my understanding of my hearing and it’s my hearing, it’s mine, no-one else’s”*. – RS (age range: 35-39 years, female, private sector)

Hearing aid users who received a more thorough explanation reported more benefits in terms of linking pure tone thresholds with relatable sounds in their everyday environments. Moreover, participants felt the process of linking audiogram results to everyday sounds gave them a better understanding of why they could not hear certain sounds in their daily life and helped them feel more confident in relaying results to communication partners. Many respondents felt they would have benefited from being given a copy of their audiogram results to monitor the progression of their hearing loss and consolidate their knowledge outside the clinic.

> *“They say there has not been much change and it’s like well actually I want more information than that because my hearing’s really important to me. It is about understanding how to interpret an audiogram for one. Oh yes, oh this is why I can’t hear these sounds that I know… It can give me more confidence*.*” -* RS (age range: 35-39 years, female, private sector)
>
> *“My graph, it’s my hearing, I always get a copy. I’ve got a copy of nearly every test I’ve done. If I’m not offered it, I get offended. I want a copy*.*”* – NO (age range: 56-60 years, female, private & public sector)
>
> *“when it was explained from my graph that the low frequencies were the ones that I was missing which is why I find men more difficult to hear often because they’ve got lower voices or deeper tones and that sort of, it was almost like a little bit of a jigsaw puzzle going oh, that’s why I can’t hear speech” –* HG (age range: 51-55 years, female, private & public sector)

### Patient-centred care

Participants shared experiences of individualised care within audiology services and recommendations for improvements in this area. A key suggestion was to include more specific, personalised questioning within the assessment, especially regarding the impact of hearing loss on employment, education, leisure, and home life, so audiologists could provide or recommend suitable intervention options.

> *“And so, the impression I get from audiologists is they think my lifestyle is just like an old person’s lifestyle and it isn’t. Every patient is different and that’s really crucial to the advice they can give as an audiologist*.*”* -RS (age range: 35-39 years, female, private & public sector)
>
> *“Just with having one conversation she really got it, straight away, she saw how upset I was and understood about the impact it has, because that’s it isn’t it, it’s not just oh you know you have a bit of trouble with the phone, it’s like well that impacts on everything. So, I think the wider issues that hearing loss brings, not just to my identity, to how I fit in with you know my relationships, going out-everything”* - ML (age range: 40-45 years, female, public sector)

Hearing aid services have high technical demands, therefore participants felt audiologists need to balance optimising hearing aid technology (e.g., performing real ear measurements) with patient communication. This could help services become more patient-focussed (rather than the technology- or clinician-focus). Other experienced hearing aid users wanted to know more about the technological capabilities of their hearing aids and additional devices that could be beneficial for managing hearing difficulties at home or at work.

> *“My audiologist made me feel that he was committed to my needs, my specific ones. With a lot of audiologists it’s about right ‘let’s give you technology’, but my audiologist knows my needs above the hearing aids are really important. And I know that if things weren’t right I could ring and he’d get me in tomorrow. You know that’s the big difference”* -LK (age range: 51-55 years, male, private sector)
>
> *“It was more about the hearing aids than the person with the hearing loss. And it shouldn’t be”*. -HG (age range: 51-55 years, female, private & public sector)

Experienced hearing aid users also commented on the impact of clinician continuity in hearing aid services. Some participants felt it was vital to see the same clinician at each appointment because they had taken time to build the rapport and trust. However, others felt that it was not so important if there was a significant amount of time between appointments.

> *“you have a relationship and I think that relationship and trust is really important. I realise it’s not always possible to see the same person, but I think if it can be done it’s important” –* LK (age range: 51-55 years, male, private sector)
>
> *“Seeing the same audiologist is hugely important, and it’s taken me a while to find the right person. I’ve been through so many audiologists” –* NO (age range: 56-60 years, female, private & public sector)
>
> *“I don’t think it makes any difference, you only see somebody once every three years you know, it’s not personal in any way” –* FE (age range: 56-60 years, female, public sector)

Finally, hearing aid users reported time constraints within the audiology consultation as a key barrier to patient-centred care.

> “*I’ve only been in one time, where someone has actually taken the time to go into detail. Where they ask how the hearing aid sounds, and really listen to the answer to suggest alternative settings. Apart from that, it’s always been once we’ve gone through the test, they ask are these settings and if any problems, give us a call. They don’t get time to get to know you*”-CD (age range: 56-60 years, male, public sector)
>
> *“I think with the public sector the biggest learning curve was to ask a lot more questions. I would say to anybody going to an audiologist-. ask them questions, tell them what you need your hearing aids for, what’s most important to you. From a private provider they actually do that because you know you’ve got your nice hour-long appointment to have a chat and develop a professional relationship. I think it’s really important*.*”* – HG (age range: 51-55 years, female, private & public sector)

## Discussion

To ensure that hearing aid services meet the needs of service users, a greater understanding of user experiences and proactive adaption to service delivery is necessary. To address this, the present study used an online focus group and a series of semi structured interviews to highlight key elements of the audiology assessment process and communication experienced hearing aid users’ value.

Adults with hearing loss have previously reported that audiologists were not in tune with their communication needs, instead using “information dumping with a script like approach” (Watermeyer et al., 2015). This results in patients having poor recall and understanding of most of the technical information relating to the nature, degree, and severity of hearing loss. Some evidence-based recommendations are available for audiologists to improve their reporting and communication skills at the initial audiology appointment (Grenness et al., 2015; Watermeyer et al., 2015), but there are currently no national or international standards. The current research adds to this body of knowledge by confirming that hearing aid users are exposed to various communication styles during audiology consultations. Many experienced hearing aid users preferred receiving clear, relatable explanations of pure tone audiometry results from their audiologists that could be easily related to real-word sounds. Despite a lack of consensus regarding if and how the audiogram should be explained to patients in detail, evidence-based tools could help audiologists discuss test results with patients, focussing on the functional impact of hearing loss (Bundesen, 2021).

Beyond audiometry, interviewees reported having minimal experience with formal aided and unaided functional hearing assessments, including the use of speech and/or background noise. Instead, it appears audiologists tend to rely on patients’ self-reports to determine hearing aid sound quality within the consultation room, thereby limiting ecological validity relating to everyday environments. Some hearing aid users felt it would be beneficial to keep track of hearing aid sound quality across a range of real scenarios (e.g., sound quality diary). However, the extended period between audiology appointments, particularly within the public sector, would mean that such feedback may not be considered in a timely manner. The use of remote care and ecological momentary assessment (a method to track a phenomena of interest in someone’s natural environment) could provide a solution to increasing responsivity to feedback in audiology and accommodating time-sensitive adjustments (Convery et al., 2020; Jenstad et al., 2021).

Hearing loss can negatively impact communication and participation in activities (Meyer et al., 2016) and thus it is not surprising that many experienced hearing aid users in the present study preferred audiologists asking personalized questions to individualise audiology assessment and rehabilitation. The World Health Organisation’s International Classification of Functioning, Disability and Health model (ICF) has been applied within the hearing healthcare context to recognise that hearing loss is not defined solely by the status of the objective bodily function, but also influenced by factors involving the individual within specific contexts (Lind et al., 2016). Implementation of self-report questionnaires and hearing aid validation measures can help promote a patient-focussed rehabilitation process (Hickson & Scarinci, 2007); however, a lack of consensus regarding the optimal outcome measures to use in audiology practice has been noted (Granberg et al., 2014). The ‘Brief ICF Core Set for Hearing Loss’ provides a minimum standard for the assessment of functioning in adults with hearing loss. Its use in clinical practice could allow for a more holistic approach to audiological assessment (Karlsson et al., 2021; van Leeuwen et al., 2020).

By definition, many patients presenting at an audiology department will have some form of hearing loss. Therefore, it is imperative audiology environments are as accessible as possible for the hearing impaired. However, a recent survey revealed that assistive communication devices were only available in 64% of audiology reception areas (Jama et al., 2020). The current research presents specific communication barriers experienced by hearing aid users including poor communication accommodations for hearing impaired patients and time constraints within public audiology services limiting adequate care and communication. Interviewees felt all audiology staff should participate in adequate and ongoing professional training regarding communication strategies and deaf awareness, as well as simulated hearing loss in order to better understand and empathize with the communication barriers patients encounter. Moving forward, it would be beneficial to involve patients in audiology service development and evaluation to improve accessibility for patients with hearing loss.

The financing and distribution of hearing aids varies globally; some countries offer universal or selective public insurance for hearing aid services (e.g., Australia, UK), whereas others rely solely on private insurance or out-of-pocket payments (e.g., USA, Japan; (Yong et al., 2019). Experienced hearing aid users who had received public sector care reported that the lack of individualised care and limited availability of tests and assessment tools were most likely due to time and resource constraints. Additional services available in the private sector are also likely to be associated with the commercial nature of private sector provision. Further evidence is needed to better understand which assessment and rehabilitation tools add value to a long-term hearing aid users’ experience of audiology services, technology uptake, and understanding of diagnostic testing results.

Previous research indicates audiologists tend to focus heavily on technological aspects, rather than considering how hearing aids address activity limitations and participation restrictions (Meyer et al., 2017). The current research also acknowledges the presence of sophisticated technical equipment within audiology services: experienced hearing aid users felt it would be beneficial to have a better balance between technology and patient factors during consultations. This can be considered in two ways. Firstly, there is a need to offset the technical diagnostic measures or hearing aid verification methods (e.g., real ear measurements) with personalized patient interaction. These beliefs are in line with a recent report indicating hearing aid users were interested in learning more about the technical process during the clinic session (Ryall et al., 2021), from which the authors provided suggestions to help audiologists make such information accessible. Secondly, participants within the present study felt individualised questioning would help audiologists better understand patients’ specific needs and expectations, with the goal of matching them to technology suitable to their lifestyle. Notably, the current study suggests that clinician continuity could help improve patient-clinician rapport and trust within hearing aid services more generally, although an association between clinician continuity and hearing aid outcomes has yet to be established (Bennett et al., 2016). Collectively, the current research aligns with an operationalised, patient-centred audiology rehabilitation model (Grenness et al., 2014), highlighting the need to focus on the patient-clinician relationship, adequately informing and involving patients in results interpretation, and individualising care.

### Strengths, limitations, and areas for future work

Because hearing loss and the onset of hearing aid use is most common in older adults (aged 65+; Barker et al., 2020), most research evaluating audiology service provision has focused on the outcomes and experiences of older adults. In contrast, one strength of the current study is the inclusion of younger hearing aid users and inclusion of participants with recent audiology service experiences in both the private and public sectors. This diversity permits a richer understanding of audiology experiences for a more heterogeneous and representative group of patients and acknowledges the needs of younger individuals with hearing loss. The diversity within participants’ service providers also helps facilitate comparison of results between countries. The implementation and detailed description of online techniques and accommodations for carrying out semi-structured interviews and focus groups with hearing aid users is also a particular strength of this research. These methods are applicable across a range of applications and such strategies will be beneficial for future studies involving special populations and/or in-person restrictions. One primary limitation of this study is that ethnicity and socio-economic status characteristics were not recorded; such factors influence access to health services and should be considered within future research. Also, within the participant group there was a skew towards more highly educated (degree level and higher) respondents.

## Conclusion

To ensure that hearing aid services meet the needs of the service users, we should explore user perspectives and proactively adapt service delivery, whilst maintaining cost effective, quality services. Within audiology, experienced hearing aid users’ value 1) comprehensive, relatable hearing assessments, 2) clear, concise, deaf aware patient-audiologist communication, 3) accessible services and 4) a personalised approach to recommend suitable technology and address patient specific aspects of hearing loss.

## Data Availability

The data that support the findings of this study are available on request from the corresponding author.

## Acknowledgements

We would like to thank to the hearing aid users that took part in the focus group and interviews and those that commented on the final manuscript. Finally, we would like to extend our thanks to Mel Ferguson and Cherilee Rutherford for assisting us during the initial planning of this study and reading this manuscript.

