## Supplemental materials for "Experienced hearing aid users’ perspectives of assessment and communication within audiology: A qualitative study using digital methods"

| Experienced hearing aid users’ perspectives of assessment and communication within audiology: A qualitative review using digital methods  **Supplementary Materials** | |
| --- | --- |
| Item 1 | Demographics table for all participants – page 2 |
| Item 2 | Topic guide for focus group and interviews – page 3-4 |
| Item 3 | Instructions for participants- page 4-5 |
| Item 4 | Sampling matrix – page 5 |

Table 1 Demographics of all participants (anonymised)

| Pseudo anonymised identifier | Gender | Age range | Hearing loss | Years of hearing aid use (range) | Time since contact with audiology | Hearing aid provider | Retired /Working | Educational level | Hours of hearing aid use/ day | Self-rated unaided  hearing  ability (1 = great difficulty, 100= no difficulty) | Self-rated aided  hearing ability  (1= is great difficulty, 100= no difficulty) | Hearing aid satisfaction  (1= very dissatisfied, 100= very satisfied) |
| --- | --- | --- | --- | --- | --- | --- | --- | --- | --- | --- | --- | --- |
| AB | M | 81-85 | Moderate | 15-20 | 1 year | Public | Retired | University degree | 16 | 11 | 89 | 88 |
| CD | M | 56-60 | Moderate | 5-9 | 2 year | Public | Working | Secondary school | 16 | 30 | 70 | 20 |
| EF | F | 46-50 | Moderate | 15-20 | 2 months | Private independent | Working | University degree | 7 | 50 | 90 | 100 |
| GH | F | 66-70 | Moderate | 5-9 | 1 week | Public | Retired | Post graduate qualification | 15 | 64 | 29 | 52 |
| IJ | M | 66-70 | Mild | 0-4 | 1 year | Public | Retired | Secondary school | 14 | 60 | 60 | 60 |
| KL | F | 66-70 | Mild | 5-9 | 5 months | Public | Retired | Post graduate qualification | 16 | 77 | 88 | 83 |
| LM | M | 71-75 | Moderate | 15-20 | 2 months | Public | Retired | Trades qualification | 18 | 30 | 79 | 85 |
| NO | F | 56-60 | Moderate | 21-35 | 2 months | Private (Independent) & public | Working | Post graduate qualification | 5-12 | 5 | 80 | 85 |
| PQ | M | 61-65 | Mild | 5-9 | 3 years | Private (National) | Working | Post graduate qualification | 16 | 76 | 96 | 85 |
| RS | F | 35-39 | Mild to profound | 31-35 | 2 months | Private (Independent) & public | Working | Trades qualification | 12-14 | 1 | 70 | 100 |
| TU | F | 81-85 | Moderate | 5-9 | 2 months | Private (National) | Retired | Diploma | 15 | 60 | 80 | 80 |
| WX | M | 56-60 | Moderate to severe | 36-40 | 2 years | Public | Working | University degree | 14 | 10 | 81 | 81 |
| YZ | M | 76-80 | Severe | 66-70 | 2 years | Private (Independent) & public | Retired | Diploma | 16 | 1 | 71 | 85 |
| QP | F | 20-24 | Mild to severe | 15-20 | 1 month | Public | Working | University degree | 14 | 52 | 80 | 90 |
| ON | F | 56-60 | Severe | 10-14 | 3 years | Private (Independent) | Working | Secondary school | 8 | 1 | 80 | 10 |
| ML | F | 40-45 | Moderate | 0-4 | 1 week | Public | Working | University degree | 12 | 6 | 70 | 85 |
| LK | M | 51-55 | Severe | 31-35 | 1 month | Private (Independent) | Working | University degree | 14 | 1 | 51 | 51 |
| JI | M | 86-90 | Mild | 0-4 | 1 month | Private (National) | Retired | Post graduate qualification | 12 | 76 | 100 | 100 |
| HG | F | 51-55 | Mild to moderate | 5-9 | 1 month | Private&Public | Retired | University degree | 12-16 | 34 | 24 | 84 |
| FE | F | 61-65 | Severe to profound | 15-20 | 1 month | Public | Working | PhD | 16 | 1 | 71 | 92 |
| DC | F | 61-65 | Moderate to severe | 21-25 | 2 years | Public | Retired | Secondary school | 15 | 24 | 68 | 60 |

| **A qualitative review of adult hearing aid users’ perspectives of audiological testing techniques**  **TOPIC GUIDE**  **Objective:** To obtain experienced hearing aid users’ perspectives of the audiological assessment process with specific focus on how they relate to everyday listening situations.  **Design:** A focus group and/or semi structured interview was carried out with a group of adult hearing aid users. A facilitator explored how audiology tests influenced their understanding and management of their hearing loss and hearing aids. The views were transcribed, and thematic analysis was used to understand key topics/themes in the data. | |
| --- | --- |
| **Main question** | **Optional probe questions** |
| **Opening questions:**  What prompted you to seek help with your hearing? |  |
| Can you describe your experiences of completing an audiology assessment (hearing test)? | What types of tests do you remember?  Tell me about the audiology assessment experience (environment/instructions/process)?  Can you describe your experiences of completing audiology tests in relation to your everyday listening /communication experiences?  Can you remember any hearing tests/assessments within previous audiology appointments that helped you understand your hearing loss? |
| If you think about the various audiology appointments you have had, how have the hearing healthcare professionals described your overall hearing ability to you? | Were there any words/terminology that you particularly remember and why?  Were you shown a graph/audiogram?  How did you relay the results to your friends and family?  How did you feel about the communication between yourself and the audiologists? |
| Could you describe the experience of being fitted with hearing aids for the first time | Once the hearing aids were put in your ears for the first time were you asked to perform any listening /hearing tests whilst wearing them? |
| Can you describe any tests, questionnaires or monitoring tools that were used to measure the benefit of or identify issues with the hearing aid fitting? | How was benefit of hearing aids monitored or measured after the hearing aid fitting? |
| As a long-term hearing aid user, what is your experience of audiology follow ups? | Have you been shown/sign posted towards any other assistive listening technology or support apart from your hearing aids?  How was your progress with technology monitored/issues identified? |
| Can you describe the communication / interactions you have experienced with audiologists? | With audiologist/reception/assisting staff?  As a hearing aid user, how do you feel about the communication / interactions you have experienced?  Are there specific areas of good practice or areas of improvement you have identified? |

**Instructions for participants**

| **What is the purpose of the study?** | In this study we would like to explore hearing aid user’s views and opinions of clinical audiology testing and communication/interaction between you and your audiologists |
| --- | --- |
| **Why have I been invited to take part?** | You have been invited to take part in this study as you are over the age of 18, have been using a hearing aid in each ear for over 2 years and have made recent contact with your audiology provider |
| **What will happen if I agree to take part (focus group)?** | - 1x one-hour focus group session will take place online, - This will take place via free videoconferencing software (Microsoft Teams or Zoom). The choice of platform will depend on all group members comfort levels and communication needs. This will be discussed with you at the next stage. - In each session, you will be talking with Bhavisha Parmar (Audiologist) and 5-6 other focus group participants via a video call. - It would be ideal to use a tablet, laptop or PC for the call so that you can see all group members within the gallery view - We will also ask to have a brief call with you before the main focus group to check your audio and video equipment. - You may switch the video feature off during the call if you prefer and we can continue discussions with audio only however, the video call function may help with the group interaction nature of the activity. - The interview will be based within a professional setting of their own home but will be using a headset to ensure confidentiality. - During the call, all participants will be asked to mute their microphones to avoid excess background noise. If you would like to take part in a particular discussion point you will be asked to unmute your microphone or raise your hand so that the interviewer can see that you wish to take part. - You are encouraged to wear your hearing aids during the call, to assist with your communication needs but if you feel a headset works well then please feel free to use that. Also, if you have a Bluetooth streaming device that is compatible with your hearing aids you are more than welcome to use that. We can test connection of the additional device in the trial call. - If you have any connection issues during the call please contact Bhavisha directly via email or text. - The full information sheet is available to give you more information about confidentiality and data protection |
| **What will happen if I agree to take part (Semi structured interview)?:** | - The project consists of 1x one-hour semi structured interview that will take place online, via free videoconferencing software (Microsoft Teams or Zoom). In each session, you will be talking with Bhavisha Parmar (Audiologist), via a video call. We might also ask to have a brief call with you before the main interview to check your audio and video equipment. - You may switch the video feature off during the call if you prefer and we can continue discussions with audio only however, the video call function may help with communication - You can choose whether we use Zoom or MS teams, based on your specific communication needs (e.g use of closed captions) - You are encouraged to wear your hearing aids during the call, to assist with your communication needs but if you feel a headset works well then please feel free to use that. Also, if you have a Bluetooth streaming device that is compatible with your hearing aids you are more than welcome to use that. We can test connection of the additional device in the trial call. - Given the 1:1 nature of the interview you can use your smartphone, tablet, laptop or PC for the interview - If you have any connection issues during the call please contact Bhavisha directly via email or text. - The full information sheet is available to give you more information about confidentiality and data protection |

**Sampling Matrix**

**Aim:**

- Equal representation of male and female
- Equal representation of over 60 and under 60 age groups
- Equal representation of work status (working or retired)
- Range of hearing loss severity
- Equal representation of private sector vs public sector experience

|  |  | % of participants |
| --- | --- | --- |
| **Sex** | Male | 43% |
|  | Female | 57% |
| **Age** | Under 60 | 47% |
|  | Over 60 | 53% |
| **Work status** | Currently working | 47% |
|  | Retired | 53% |
| **Hearing loss severity** | Mild | 19% |
|  | Moderate | 48% |
|  | Severe-profound | 33% |
| **Sector** | Public | 52% |
|  | Private | 29% |
|  | Public & private | 19% |

Table 2 Sampling matrix
